# OpenSAFELY: Risks of COVID-19 hospital admission and death for people with learning disabilities - a cohort study

**DOI:** 10.1101/2021.03.08.21253112

**Authors:** The OpenSAFELY Collaborative, Elizabeth J Williamson, Helen I McDonald, Krishnan Bhaskaran, Alex J Walker, Sebastian Bacon, Simon Davy, Anna Schultze, Laurie Tomlinson, Chris Bates, Mary Ramsay, Helen J Curtis, Harriet Forbes, Kevin Wing, Caroline Minassian, John Tazare, Caroline E Morton, Emily Nightingale, Amir Mehrkar, Dave Evans, Peter Inglesby, Brian MacKenna, Jonathan Cockburn, Christopher T Rentsch, Rohini Mathur, Angel YS Wong, Rosalind M Eggo, Will Hulme, Richard Croker, John Parry, Frank Hester, Sam Harper, Ian J Douglas, Stephen JW Evans, Liam Smeeth, Ben Goldacre, Hannah Kuper

## Abstract

**Objectives:** To assess the association between learning disability and risk of hospitalisation and mortality from COVID-19 in England among adults and children.

**Design:** Working on behalf of NHS England, two cohort studies using patient-level data for >17 million people from primary care electronic health records were linked with death data from the Office for National Statistics and hospitalization data from NHS Secondary Uses Service using the OpenSAFELY platform.

**Setting:** General practices in England which use TPP software.

**Participants:** Participants were males and females, aged up to 105 years, from two cohorts: (1) wave 1, registered with a TPP practice as of 1^st^ March 2020 and followed until 31^st^ August, 2020; (2) wave 2 registered 1^st^ September 2020 and followed until 31^st^ December 2020 (for admissions) or 8^th^ February 2021 (for deaths). The main exposure group was people included on a general practice learning disability register (LDR), with a subgroup of people classified as having profound or severe learning disability. We also identified patients with Down syndrome and cerebral palsy (whether or not on the learning disability register).

**Main outcome measures:** (i) COVID-19 related death, (ii) COVID-19 related hospitalisation. Non-COVID-19 related death was also explored.

**Results:** In wave 1, of 14,301,415 included individuals aged 16 and over, 90,095 (0.63%) were identified as being on the LDR. 30,173 COVID-related hospital admissions, 13,919 COVID-19 related deaths and 69,803 non-COVID deaths occurred; of which 538 (1.8%), 221 (1.6%) and 596 (0.85%) were among individuals on the LDR, respectively. In wave 2, 27,611 COVID-related hospital admissions, 17,933 COVID-19 related deaths and 54,171 non-COVID deaths occurred; of which 383 (1.4%), 260 (1.4%) and 470 (0.87%) were among individuals on the LDR. Wave 1 hazard ratios for individuals on the LDR, adjusted for age, sex, ethnicity and geographical location, were 5.3 (95% confidence interval (CI) 4.9, 5.8) for COVID-19 related hospital admissions and 8.2 (95% CI: 7.1, 9.4) for COVID-19 related death. Wave 2 produced similar estimates. Associations were stronger among those classed as severe-profound and among those in residential care. Down syndrome and cerebral palsy were associated with increased hazard of both events in both waves; Down syndrome to a much greater extent. Hazards of non-COVID-19 related death followed similar patterns with weaker associations.

**Conclusions:** People with learning disabilities have markedly increased risks of hospitalisation and mortality from COVID-19. This raised risk is over and above that seen for non-COVID causes of death. Ensuring prompt access to Covid-19 testing and health care and consideration of prioritisation for COVID-19 vaccination and other targeted preventive measures are warranted.

## Background

Identification of high-risk groups for severe outcomes from COVID-19 is critically important in risk stratification, including informing vaccine prioritisation and other targeted preventive measures. People with learning disability, who total more than one million people in England alone or 2% of the adult population, may be one such vulnerable group^1^. As of February 2021, the Learning from Death Reviews (LeDeR) programme reported that 1405 people with a learning disability had died from COVID-19 in England since February 2020^2^. The true number is likely to be far higher due to gaps in registration of learning disability. Emerging evidence from the first wave of the COVID-19 epidemic showed that people with learning disability were at higher risk from mortality^3–7^ compared to others in the population. For instance, the Oxford RCGP Research and Surveillance Centre (RSC) sentinel network reported an odds ratio of 1.96 (95% confidence interval (CI) 1.22 to 3.18) for mortality during the first wave of infection in the UK, among people with learning disability compared to those without.^4^ People with Down syndrome may be at particularly high risk; an analysis of primary care data from 8 million adults reported a hazard ratio of 10.4 (95% CI 7.1 to 15.2) for COVID death associated with Down syndrome.^8^ However, existing studies on the association of learning disability with COVID-19 severe outcomes do not include the second wave of the pandemic, and frequently adjusted for variables which might be on the causal pathway, such as deprivation and comorbidities, complicating interpretation of the results^4^. There is also lack of clarity on the elevated risk of COVID-19 deaths among individuals with milder learning disability, and this needs exploration.^9^

The higher risks of premature death among people with learning disability in England is well-known,^1,10^ and triggered the establishment of GP Learning Disability registers to allow for better provision of their healthcare. There are a number of mechanisms by which their risk of COVID-19 mortality may also be greater. People with learning disability have a higher prevalence of COVID-19 mortality risk factors, including obesity, diabetes, epilepsy and poverty.^11–13^ Medical conditions underlying the learning disability may confer additional risk; for instance, people with Down syndrome are more vulnerable to immune dysregulation, congenital heart disease and respiratory conditions^14^. Many people with learning disability in England live in residential care or receive community based social care, and therefore have frequent contact with carers and other care recipients and face challenges in physical distancing. Difficulties understanding protective measures needed, compounded by lack of accessible information, further increases their vulnerability to infection. Healthcare access and quality, including prevention and treatment, is frequently worse for people with learning disability, and leads to avoidable deaths.^10^ Treatment failures^10^, including do not resuscitate orders^15^, may increase their risk of death once infected.

Until February 24, 2021, the national recommendations for prioritisation of COVID-19 vaccination in England included all adults with cerebral palsy, severe or profound learning disability, Down syndrome, and the whole resident population in care settings where a high proportion of residents would be eligible for vaccination (for example due to learning disability).^16,17^ Consequently, not everyone on the Learning Disability registers was eligible for COVID-19 vaccination, in particular people with mild to moderate learning disability from causes other than Down syndrome or cerebral palsy who are not living in residential care.

This work was undertaken rapidly in response to an urgent need to inform policy-making on vaccination prioritisation in the UK and elsewhere.

The aim of this study is to use linked electronic health records within the OpenSAFELY platform to rapidly describe the risk of COVID-19 related hospitalisation and mortality among children and adults in England with learning disability compared to the general population. A subsidiary aim is to separate the risk by types of learning disability (severe-profound; cerebral palsy, Down syndrome, learning disability register), including people with learning disability not originally included in the first 6 priority groups of the Phase 1 vaccination priority list in the UK.

## Methods

### Study design

Two population-based observational cohort studies of patients in England were performed using data within the OpenSAFELY platform.

### Data

We used data from primary care linked to secondary care and mortality records in England. Records were linked to the NHS England inpatient activity data sets from Secondary Uses Service (SUS) data extracts including data from inpatient activity data sets for ascertaining COVID-related hospitalisations^18^; and Office for National Statistics (ONS) death data for ascertaining COVID-19 related deaths. The dataset analysed within OpenSAFELY is based on 24 million people currently registered with GP surgeries using TPP SystmOne software, approximately 40% of the population in England. All data is pseudonymized and includes coded diagnoses (using Read version 3, CTV3 codes), medications and physiological parameters. No free text data are included.

### Study population

The first cohort comprised patients (males and females, aged up to 105 years) registered as of 1^st^ March 2020 in a general practice which employs the TPP system and followed until 31^st^ August, 2020. Patients with missing age or a recorded age over 105 years, missing gender, or missing postcode (from which much of the household and geographic information is calculated) were excluded. The second cohort were similarly defined, but included patients registered as of 1^st^ September 2020 in a general practice which employs the TPP system and followed until 31^st^ December 2020 (admissions) or 8^th^ February 2021 (deaths). Membership of the two cohorts may differ slightly due to patients leaving and joining TPP practices and to patients dying prior to the second cohort. These two time periods correspond to the two main “waves” of COVID-19 infection experienced in England during 2020.^19^ In particular, the 1st September 2020 had the lowest number of COVID-19 related deaths since the start of the pandemic.^20^

#### Exposures

All codelists used to define exposure groups are provided online, with links given in the Supplementary Materials. The main exposure group was individuals on the learning disability register. This register contains a subset of individuals with learning disability; it is not a comprehensive list. However, the register provides a simple and practical means of identifying people for vaccine prioritisation or implementation of other public health measures. A subset of the codes used to define the learning disability register classified the learning disability as severe or profound, and were used to class a subset of individuals as having severe or profound learning disability.

Due to the absence of a comprehensive indicator of residential care, individuals living in a household containing at least five individuals identified as being on the learning disability register were classed as being in residential care. Households were identified based on GP registered addresses as of 1st February 2020, standardised and corrected using publicly available house sale data to remove registrations which are likely not current. We use the term “residential care” throughout, although we note that this includes a range of settings (care homes, educational settings, sheltered accommodation, etc.) and there is likely to be misclassification.

Individuals with Down syndrome and cerebral palsy were identified based on GP codes (details in Supplementary Materials).

### Outcomes

The outcomes for this study are (i) COVID-19 related death (defined as a COVID-19 ICD-10 code of U07.1 or U07.2 anywhere on the death certificate - ascertained from ONS death certificate data)) and (ii) COVID-19 related hospitalisation (defined as admissions with any ICD-10 admission diagnosis (not restricted to primary diagnosis) of U07.1 or U07.2 - ascertained from SUS data). Individuals who experienced a COVID-19 related hospitalisation and then a death contributed to both outcomes. An additional outcome of non-COVID-19 related death was also considered (ascertained from ONS death certificate data, excluding deaths classed as COVID-19 related).

#### Covariates

Covariates included demographics (age, sex, ethnicity and geographical area), which could act as potential confounders, and deprivation (index of multiple deprivation) as a potential confounder and/or mediator. To consider mediation by physical comorbidities that are also indications for vaccination we included body mass index > 40 kg/m2, chronic cardiac disease, atrial fibrillation, deep vein thrombosis/pulmonary embolism, diabetes (further grouped by level of control, as measured by the latest HbA1c measurement), chronic liver disease, stroke, transient ischaemic attack, dementia, asthma requiring use of oral corticosteroids, other chronic respiratory disease, reduced kidney function, dialysis, organ transplant, asplenia, other conditions leading to immunosuppression, and haematological cancer. We also included non-haematological cancer diagnosed in the last year, rheumatoid arthritis/lupus/psoriasis, and inflammatory bowel disease as common indications for immunosuppressing medication. These aim to map to the existing physical indications for vaccination among 16-64 year olds in England; however data on epilepsy was not available for this analysis. To exclude individuals already prioritised for vaccination we additionally ascertained other neurological conditions and serious mental illness. These measures were obtained from medical records (details in Supplementary Materials).

### Statistical methods

Analysis was undertaken separately for adults 16 years and over and children under 16. The three analysis steps below were repeated for the following exposures: being on the learning disability register (all, then divided into severe-profound versus mild-moderate and residential care versus non-residential care), Down syndrome and cerebral palsy.

The two cohorts, for each wave, were analysed separately. Analysis used Cox proportional hazards model for (i) COVID-19 related mortality and (ii) COVID-19 related hospital admission, stratified by local geographical area, as measured by the Sustainability and Transformation Partnership, to account for differing patterns of infection over time in different regions, with days in study as the timescale. Follow-up was censored at competing events (non-COVID death for mortality analyses and any death for hospital admission analyses) to target the cause-specific hazard. Models adjusted for confounders (age, sex, ethnicity), additionally for deprivation, residential care status and physical comorbidities (described above), and then adjusted for all of these factors simultaneously. Exposure interactions with broad age strata (16-<65, 65-<75, 75+) were explored.

Similar Cox models were fitted for COVID-19 related hospital admission, after excluding individuals who were already prioritised for vaccination due to age or comorbidities, as part of the first 6 priority groups of Phase I in the UK. These were individuals 65 years and over and individuals with codes for physical conditions indicating priority for vaccination described above, other neurological conditions, severe mental illness, Down syndrome, cerebral palsy, and severe-profound learning disability. These Cox models adjusted for the confounders, then separately for deprivation and residential care status.

Finally, Cox models for non-COVID-19 related death were fitted, adjusting for the same variables as previous models.

For children under 16 years, these analyses were undertaken separately, omitting analyses looking at COVID-19 related death and full adjustment for comorbidities due to much smaller numbers of outcomes.

#### Missing data

A complete case approach was taken to missing ethnicity data (∼25% of records). Previous analyses in these data suggest the assumption required for complete case analysis - that missingness is unrelated to outcome given covariates - are approximately satisfied here^12^. Individuals with missing BMI were assumed to be non-obese. Patients with no serum creatinine measurement were included in the “no evidence of poor kidney function”. Patients with diabetes but no Hba1c measurement were included in a separate “diabetes, no Hba1c” category.

## Software and Reproducibility

The pre-specified study protocol is archived with version control https://github.com/opensafely/Published-Protocols/blob/master/Learning_Disability_Covid_Protocol_2021_10_02.pdf)

## Patient and Public Involvement

We have developed a publicly available website https://opensafely.org/ through which we invite any patient or member of the public to contact us regarding this study or the broader OpenSAFELY project.

## Results

Table 1 shows the baseline characteristics of individuals aged 16 and over included in the analysis and Table 2 shows characteristics of those under 16 years old (characteristics in wave 2 were very similar).

Among 14,301,415 included individuals aged 16 or over, 90,095 (0.63%) were identified as being on the learning disability register (Table 1). Of these, 16,115 (18%) were identified as having severe-profound learning disability, and 8,027 (9%) as being in residential care. 8,022 individuals were identified as having Down syndrome, of whom 7124 (89%) were on the learning disability register. 18,264 individuals were identified as having cerebral palsy, of whom 6,913 (38%) were on the learning disability register. Those on the learning disability register were more likely to be male, younger and living in more deprived areas. Comorbidities were fairly similar across groups, with more diabetes, obesity, other neurological disease, and diagnoses of serious mental illness among those on the learning disability register.

**Table 1.**
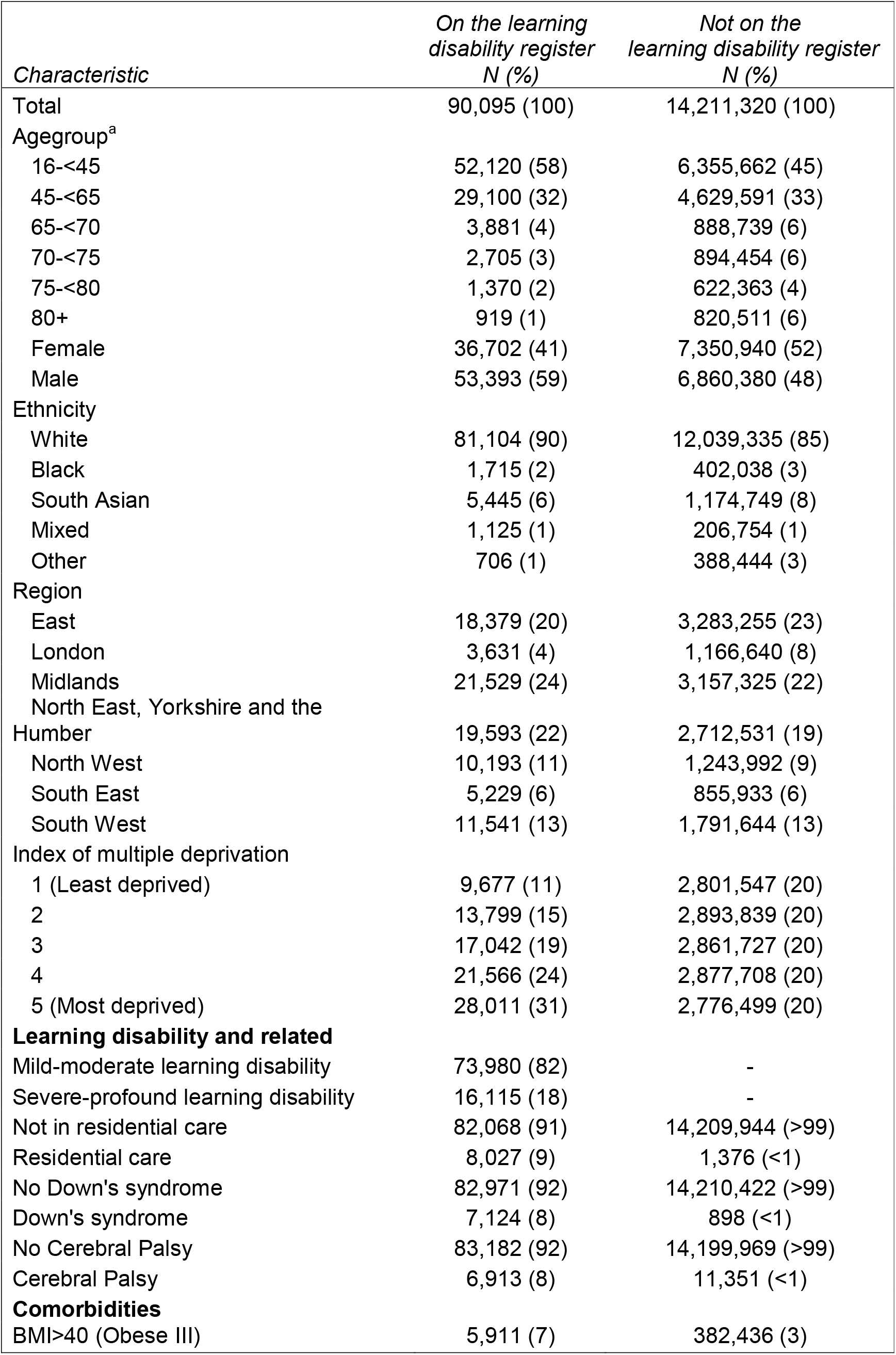
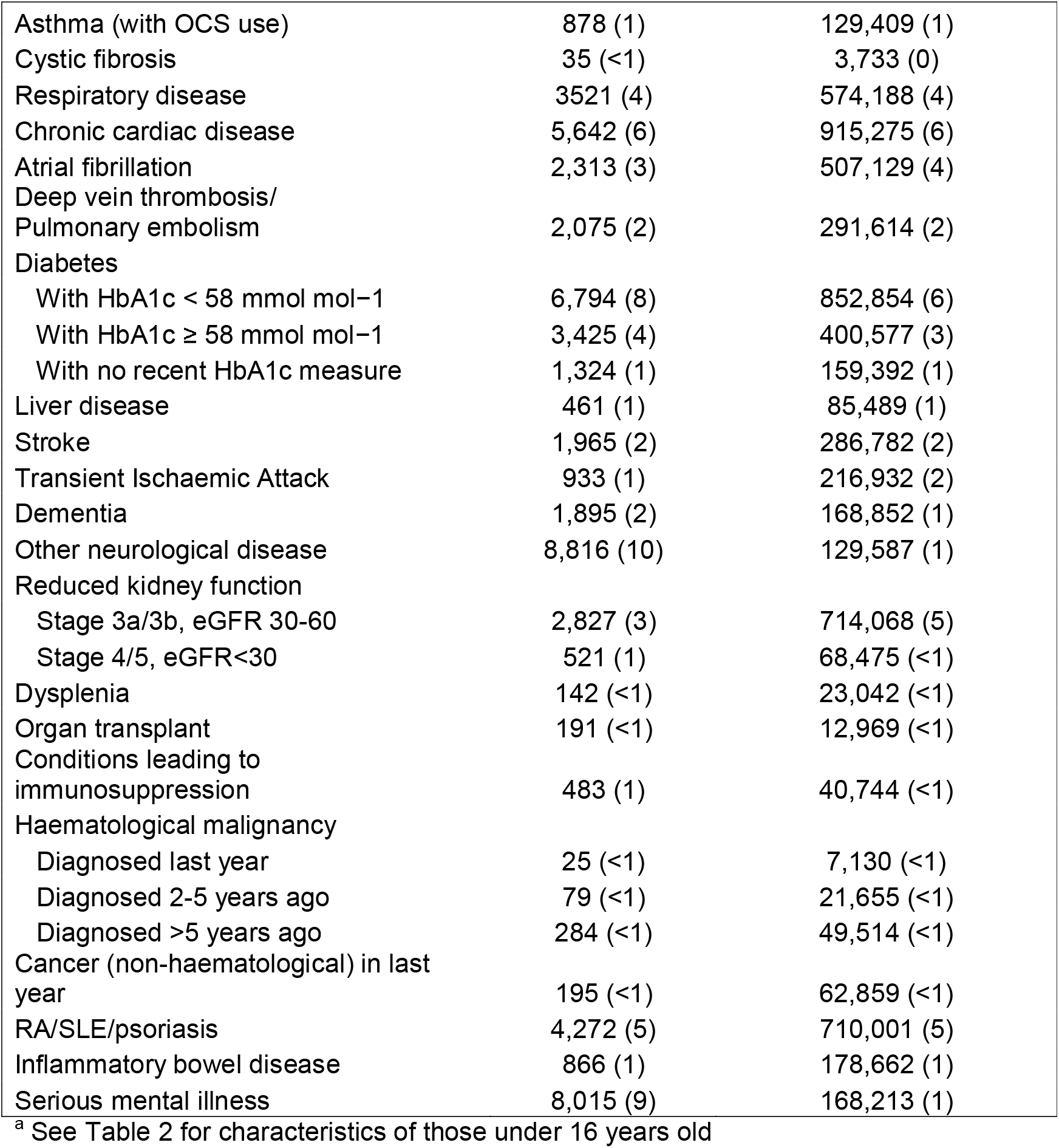
Baseline characteristics of individuals 16 years and older

**Table 2.** Baseline characteristics of individuals under 16 years old

| *Characteristic* | *On the learning*<br>*disability register*<br>*N (%)* | *Not on the*<br>*learning disability register*<br>*N (%)* |
| --- | --- | --- |
| Total | 9,114 (100) | 2,615,239 (100) |
| Female | 2,965 (33) | 1,275,295 (49) |
| Male | 6,149 (67) | 1,339,944 (51) |
| Ethnicity |  |  |
| White | 6,971 (76) | 2,070,441 (79) |
| Black | 369 (4) | 96,159 (4) |
| South Asian | 1,256 (14) | 277,942 (11) |
| Mixed | 299 (3) | 99,477 (4) |
| Other | 219 (2) | 71,220 (3) |
| Region |  |  |
| East | 1,743 (19) | 618,596 (24) |
| London | 552 (6) | 167,325 (6) |
| Midlands | 2,188 (24) | 608,128 (23) |
| North East, Yorkshire and the<br>Humber | 2,050 (22) | 493,649 (19) |
| North West | 966 (11) | 224,103 (9) |
| South East | 566 (6) | 169,577 (6) |
| South West | 1,049 (12) | 333,861 (13) |
| Index of multiple deprivation |  |  |
| 1 (Least deprived) | 1,215 (13) | 485,858 (19) |
| 2 | 1,383 (15) | 471,778 (18) |
| 3 | 1,584 (17) | 483,503 (18) |
| 4 | 2,069 (23) | 533,144 (20) |
| 5 (Most deprived) | 2,863 (31) | 640,956 (25) |
| **Learning disability and related** |  |  |
| Mild-moderate learning disability | 7,740 (85) | - |
| Severe-profound learning disability | 1,374 (15) | - |
| Not in residential care | 9,054 (99) | 2,615,192 (>99) |
| Residential care | 60 (1) | 47 (>1) |
| No Down's syndrome | 8,207 (90) | 2,613,511 (>99) |
| Down's syndrome | 907 (10) | 1,728 (<1) |
| No Cerebral Palsy | 8,614 (95) | 2,611,120 (>99) |
| Cerebral Palsy | 500 (5) | 4,119 (<1) |
| **Comorbidities** |  |  |
| BMI>40 (Obese III) | 70 (1) | 4408 (<1) |

Among 2,624,353 included individuals under 16 years (Table 2), 9,114 (0.35%) were identified as being on the learning disability register. Of the 2,635 individuals identified as having Down syndrome, only 907 (34%) were identified as being on the learning disability register. Of the 4,619 individuals identified as having cerebral palsy, 500 (11%) were identified as being on the learning disability register.

### COVID-19 related hospital admissions and deaths

Among individuals 16 years and older during wave 1 (1 March 2020 - 31 August 2020; 183 days), 30,173 COVID-related hospital admissions occurred, 538 (1.8%) among individuals on the learning disability register; 13,919 COVID-19 related deaths occured, 221 (1.6%) among individuals on the learning disability register (Table 3). 69,803 non-COVID deaths occurred, 596 (0.85%) among individuals on the learning disability register.

**Table 3.** Estimated hazard ratios for COVID-19 outcomes among adults 16 years and over, adjusted for potential confounders (age, sex, ethnicity, geographical region)

| *Exposure category* | *Wave 1 (1 Mar 2020 – 31 Aug 2020)* |  |  |  | *Wave 2 (1 Sept 2020 – Nov 2020 (admissions) or Jan 2021 (death))* |  |  |  |
| --- | --- | --- | --- | --- | --- | --- | --- | --- |
|  | *COVID-19 hospital admission* |  | *COVID-19 related death* |  | *COVID-19 hospital admission* |  | *COVID-19 related death* |  |
|  | *Events* | *HR (95% CI)* | *Events* | *HR (95% CI)* | *Events* | *HR (95% CI)* | *Events* | *HR (95% CI)* |
| LDR |  |  |  |  |  |  |  |  |
| No | 29,635 |  | 13,698 |  | 27,228 |  | 17,673 |  |
| Yes | 538 | 5.33 (4.88, 5.83) | 221 | 8.18 (7.12, 9.39) | 383 | 3.64 (3.28, 4.03) | 260 | 7.39 (6.52, 8.38) |
| by severity: |  |  |  |  |  |  |  |  |
| Mild-moderate | 391 | 4.76 (4.32, 5.25) | 158 | 7.1 (6.08, 8.3) | 281 | 3.27 (2.91, 3.67) | 179 | 6.2 (5.37, 7.17) |
| Severe-profound | 147 | 7.8 (6.47, 9.4) | 63 | 13.21 (9.99, 17.48) | 102 | 5.27 (4.31, 6.44) | 81 | 12.81 (10.25, 16) |
| by residential care status: |  |  |  |  |  |  |  |  |
| Not in residential care | 438 | 4.89 (4.46, 5.37) | 182 | 7.82 (6.77, 9.02) | 318 | 3.36 (2.99, 3.76) | 211 | 6.92 (6.04, 7.94) |
| In residential care | 100 | 8.77 (6.8, 11.3) | 39 | 10.42 (6.88, 15.78) | 65 | 6.14 (4.58, 8.22) | 49 | 10.41 (7.51, 14.43) |
| Down's syndrome |  |  |  |  |  |  |  |  |
| No | 30,098 |  | 13,878 |  | 27,558 |  | 17,868 |  |
| Yes | 75 | 10.56 (8.45, 13.2) | 41 | 35.96 (26.38, 49.02) | 53 | 7.22 (5.52, 9.46) | 65 | 42.91 (33.54, 54.9) |
| Cerebral Palsy |  |  |  |  |  |  |  |  |
| No | 30,076 |  | 13,889 |  | 27,536 |  | 17,902 |  |
| Yes | 97 | 4.93 (3.84, 6.34) | 30 | 5.86 (4.14, 8.3) | 75 | 3.7 (2.93, 4.69) | 31 | 4.54 (3.19, 6.47) |

LDR Learning Disability Register; HR Hazard Ratio; CI confidence interval

In wave 2 (1 September 2020 - 31 December 2020 (admissions) or 8 February 2021 (death); 121 and 160 days, respectively), 27,611 COVID-related hospital admissions occurred, 383 (1.4%) among individuals on the learning disability register; 17,933 COVID-19 related deaths occured, 260 (1.4%) among individuals on the learning disability register. 54,171 non-COVID deaths occurred, 470 (0.87%) among individuals on the learning disability register.

Figure 1 shows cumulative COVID-19 related mortality and hospital admissions during the study period, accounting for sex, age and ethnicity for individuals on the learning disability register and those not. Both graphs show a clear increase in events among those on the learning disability register, with a flattening off apparent during the period between waves of infection.

**Figure 1.**
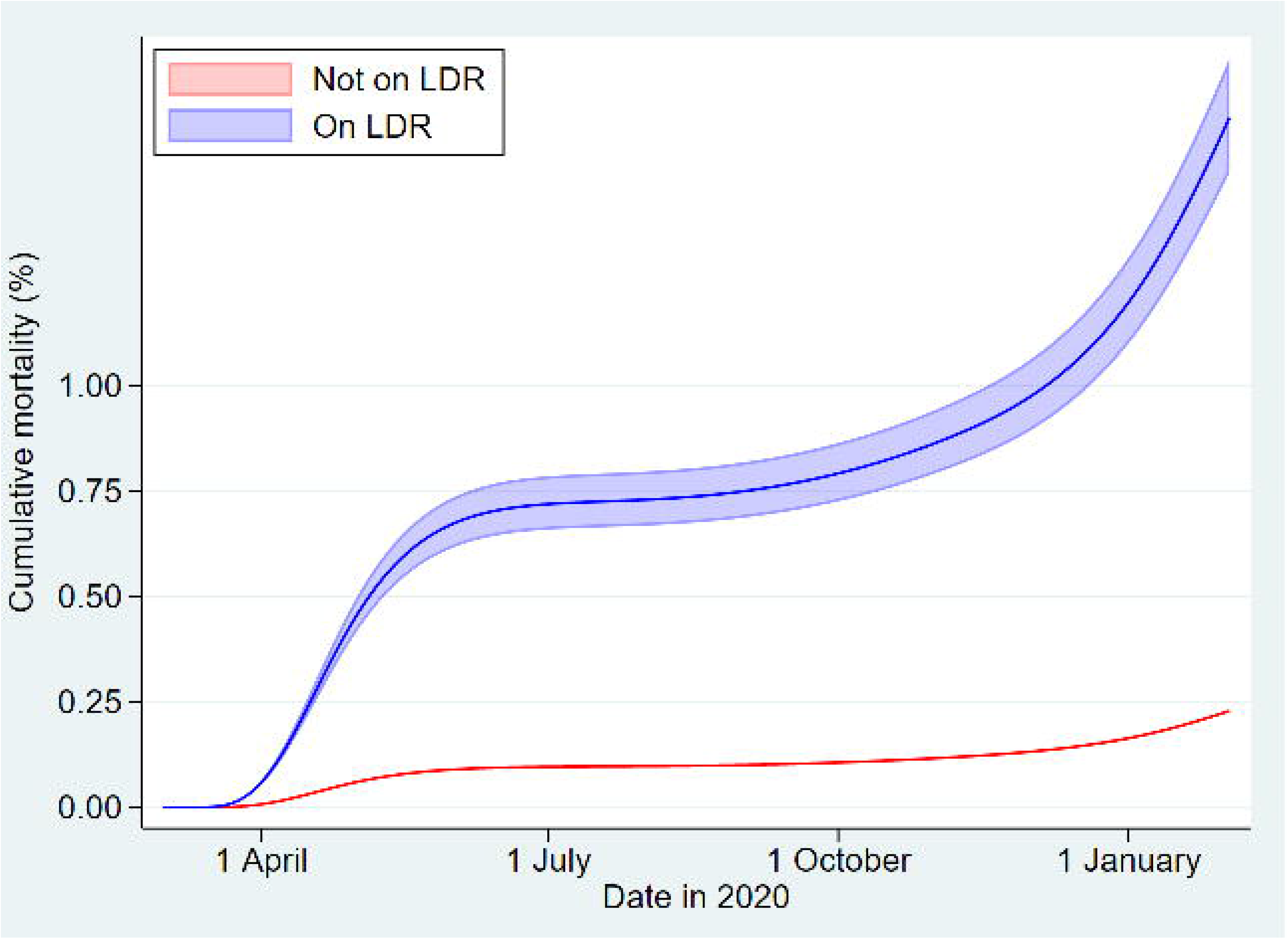
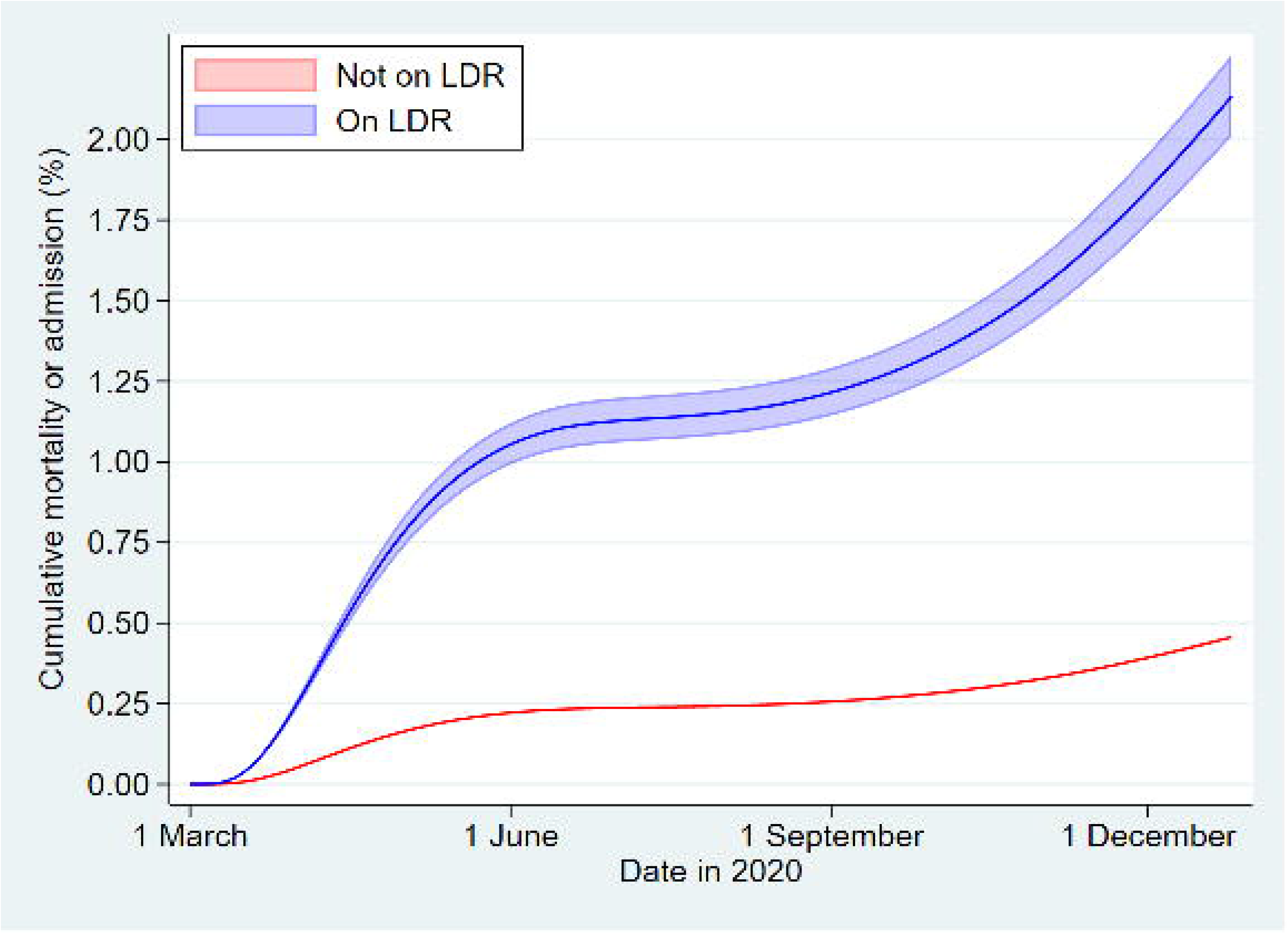
Cumulative (a) COVID-19 mortality and (b) COVID-19 hospital admission or mortality for individuals on the learning disability register and not with 95% CI, standardised by age, sex and ethnicity, among individuals 16 years and above through the two waves o infection.

Among individuals under 16 years of age, 281 COVID-19 related hospital admissions occurred in wave 1 and 292 in wave 2. These numbers cannot be split by learning disability status due to stringent redaction rules applied to protect patient privacy.

### Hazard ratios for COVID-19 hospital admission and death

Adjusted for age, sex, ethnicity and geographical location, the estimated hazard ratio for COVID-19 related hospital admission in wave 1 was 5.3 (95% confidence interval (CI) 4.9, 5.8) for individuals on the learning disability register (Table 3). For COVID-19 related death, the hazard ratio was 8.2 (95% CI: 7.1, 9.4). Wave 2 produced similar estimates (HR for COVID-19 related hospital admission 3.6 (95% CI 3.3, 4.0), HR for COVID-19 related death 7.4 (95% CI 6.5, 8.4)). These associations were stronger among those classed as severe-profound and among those in residential care. Down syndrome was associated with greatly increased hazard of both events in both waves, (wave 1 HR for COVID-19 related hospital admission 10.6 (95% CI 8.5, 13.2), wave 1 HR for COVID-19 related death 36.0 (95% CI 26.4, 49.0; similar numbers for wave 2). Cerebral palsy was associated with higher hazards to a lesser extent (wave 1 HR for COVID-19 related hospital admission 4.9 (95% CI 3.8, 6.3), wave 1 HR for COVID-19 related death 5.9 (95% CI 4.1, 8.3; similar numbers for wave 2).

Further adjustment for deprivation, residential care and physical comorbidities only slightly attenuated these associations (Supplementary tables). The confounder-adjusted hazard ratio for wave 1 COVID-19 related hospital admission for being on the learning disability register, HR 5.3 (95% CI 4.9, 5.8), reduced to 3.9 (95% CI 3.6, 4.3) after full adjustment. In Down syndrome, however, adjustment for both residential care and comorbidities greatly attenuated hazard ratios (for example, the wave 1 hazard ratio for COVID-19 related hospital admission reduced from 10.6 (95% CI 8.5, 13.2) to 6.3 (95% CI 4.9, 8.2) after adjustment for residential care and 7.2 (95% CI 5.8, 9.0) after adjustment for physical comorbidities and 4.64 (95% CI 3.6, 6.0) after adjusted for all.

### Hazard ratios for COVID-19 hospital admission among those under 16

Among individuals under 16 years old, being on the learning disability register was associated with increased hazard of COVID-19 related hospital admissions (wave 1 HR 5.1 (95% CI 1.9, 14.0), wave 2 HR 8.8 (95% CI 4.9, 15.8), Supplementary Tables).

### Effect modification by age

In wave 1, an interaction was observed between being on the learning disability register and age, on the hazard of COVID-19 related death, with those 16-<65 having a hazard ratio 12.3 (95% CI 10.0, 15.1), 65-<75 HR 10.1 (8.3, 13.3) and 75+ HR 4.2 (95% CI 3.2, 5.5), with similar estimates seen for wave 2. A similar interaction was not observed for COVID-19 related hospital admission in either wave.

### Hazard ratios among individuals not prioritised for vaccination

After excluding people 65 years and older and those with comorbidities listed above, the estimated hazard ratio of COVID-19 related hospital admission was 6.7 (95% CI 1.6, 28.2) after adjustment for age, sex, ethnicity and geographical location, with little change after adjustment for deprivation or residential care status. Too few events were observed to analyse finer groupings or wave 2 events.

### Hazard ratios for non-COVID-19 death

Adjusted for age, sex, ethnicity and geographical location, the estimated hazard ratio for non-COVID-19 related death was 3.7 (95% CI 3.4, 4.0) in wave 1 and 3.9 (3.6, 4.) in wave 2. Associations were stronger among those classed as having severe-profound learning disability. Associations were slightly stronger among those classed as being in residential care. Down syndrome was associated with a greatly increased hazard of non-COVID-19 related death (wave 1 HR 12.3 (95% CI 10.0, 15.2; similar numbers for wave 2). Cerebral palsy was associated with higher hazards to a lesser extent (wave 1 HR for COVID-19 related death 3.2 (95% CI 2.6, 4.0; similar numbers for wave 2).

## Discussion

Our data show a higher risk of hospitalisation and deaths for all the groups with learning disability relative to comparators. Generally, the pattern of hazard ratios is consistent for waves 1 and 2, and hospitalisations and deaths. Hazard ratios appear slightly lower for wave 2 than wave 1 for both hospitalisations and deaths. Higher risks were observed among those with severe-profound learning disability compared to mild-moderate, which was not explained by measured physical comorbidities or residential care status. However, the absolute number of deaths was higher among people with mild-moderate learning disability. For Down Syndrome and cerebral palsy, we observed higher risk among those on the learning disability register. Higher risk remains among those on the learning disability register who do not have Down syndrome or cerebral palsy, compared to the general population. This is not explained by measured physical comorbidities or residential care status. After excluding people who were prioritised for vaccination among groups 1-6 of Phase I in England due to reasons of age or comorbidity, those on the learning disability register had a remaining substantial increased risk of COVID-19 related hospital admission. Higher risks of non-COVID-19 related deaths were also observed in people with learning disabilities, though associations were much less strong than for COVID-19 death; this is in contrast with most other risk factors which appear to have a similar magnitude of association with both COVID-19 related and non-COVID-19 related death^21^.

### Findings in context

Our findings are consistent with the existing literature. The Oxford RCGP Research and Surveillance Centre sentinel network, including 4.4 million people nationally representative from England, reported two-fold higher mortality among people with learning disability (OR 1.96, 95% CI 1.22, 3.18, p<0.01) after extensive adjustment^4^. Public Health England used different sources of data and estimated that, up to June 2020, people with learning disability had approximately 6.3 times the mortality rate as the general population^22^. Overall, COVID-19 was responsible for at least half of deaths among people with learning disability during this period. Data from Scotland also showed that adults with intellectual disabilities had higher rates of COVID-19 infection, severe infection and mortality^5^. Elevated risks remained after adjusting for age, sex and deprivation (Standardised severe infection ratio: 2.59, 95% CI 1.80, 3.39; Standardised mortality ratio 3.20 95% CI 2.1, 4.25). Higher mortality rate among people with learning disability has also been demonstrated in Wales^6^, New York State^7^, and the USA more broadly^3^. Not only are death rates higher for people with learning disability, but deaths occur at younger ages^3,8,9,22^. Indeed, the particularly elevated mortality rates among younger people with learning disability noted in our study have also been reported in the USA^3^.

The particularly high risk for people with Down Syndrome was demonstrated for the first wave by Clift and colleagues using the QResearch population level primary care database^8^. In their cohort of 8 million adults in England from January to June 2020, the age- and sex-adjusted hazard ratio for COVID-19 death for adults with versus without Down syndrome was 24.94 (95% CI 17.08, 36.44), which reduced to 10.39 (95% CI 7.08, 15.23) after extensive adjustment (deprivation, BMI, cardiovascular, pulmonary and other disease, residential status, ethnicity). Data for cerebral palsy is more limited, but the QResearch analyses showed a higher mortality rate for this group (2.66, 95% CI 1.62, 4.36). Clift and colleagues obtained a fully adjusted hazard ratio of 1.27 (95% CI 1.16, 1.40) for COVID-19 death in those with learning disabilities other than Down syndrome, in contrast to our higher estimates. This discrepancy is likely due to differences in adjustment. We chose not to adjust for many comorbidities, viewing many of these as consequences of the learning disability thus part of the causal pathway. In our analyses, children with learning disability had a higher risk of hospital admission for COVID-19, notwithstanding the extremely small numbers. Similarly, existing studies also indicate that children with learning disability are more vulnerable to requiring hospitalization and critical care due to COVID-19 outcomes^23–26^. Evidence on mortality remains lacking for children.

The hazard rates for COVID-19 outcomes in our study attenuated after adjustment for deprivation. Residential status also partially explained the higher risks of severe COVID-19 outcomes in people with learning disability. However, residential care for people with learning disability may not raise risk as much as in other care settings, perhaps because there are generally fewer beds^22^ in these facilities. An important driver appears to be comorbidities, reflecting the higher prevalence of these COVID-19 risk factors among people with learning disability^11,13^. However, large excess mortality rates remained after extensive adjustment, as is also apparent in previous studies^8^. This pattern implies that other drivers may be relevant, including inherent clinical vulnerabilities for people with certain conditions and concerns about healthcare quality, as also indicated by the higher case fatality rates among people with learning disability (Scotland: 30% versus 24%^5^; New York: 15,0% versus 7.9%^7^; USA 18-74 year olds: 4.5% versus 2.7%^3^).

### Strengths and weaknesses

Key strengths and weaknesses of the OpenSafely platform have been outlined previously^12^. An important strength for the current analyses is that the study is large, including records of approximately 40% of the English population, allowing disaggregation by learning disability grouping. We had comprehensive data on participants from medical records, allowing us to adjust analyses successively to explore mechanisms for the association of learning disability and adverse COVID-19 outcomes. Furthermore, we were able to assess excess risks in both waves 1 and 2 of the COVID-19 epidemic, in terms of both hospitalization and mortality outcomes. We considered children as well as adults.

There are also important limitations. It is not possible to identify everyone with a learning disability from medical records alone, which may have under-estimated hazard ratios. For instance, the most recent data, from 2015, suggests that 23% of people with learning disability are included on the registers^1^. Our hospitalisation data included only completed hospital admissions, thus will have under-ascertained this outcome towards the end of wave 2. Data was not available on epilepsy, which is both a COVID-19 risk factor and more common among people with learning disability^11,13^. We also had an incomplete measure of residential care. There was missing data, in particular for ethnicity, although we do not anticipate that this had a meaningful impact on the results. We did not have data on quality of treatment and so were not able to explore all our hypothesised pathways between learning disability and adverse COVID-19 outcomes. Furthermore, these analyses focussed only on severe COVID-19 outcomes, and did not explore impacts on physical and mental health of people with learning disability which are likely to occur as a result of lockdown and other restrictions^23^ and require mitigation. We focused on the general population, so our analyses explore the combination of infection and severe outcomes once infected, thus we are unable to disentangle associations with those two steps in the process. However, focusing on the infected only would induce biases due to unrepresentative testing.

### Policy implications and interpretation

In February, 2021, the Joint Committee on Vaccination and Immunisation updated its guidance^27^ to include everyone on the Learning disability registers, as well as people with Down Syndrome, cerebral palsy or in residential care, as priority groups to receive the vaccine. This change was informed by a prior version of this analysis, demonstrating the increased risk in the remaining group. The current Learning Disability registers are incomplete^1^, however, and updating them will facilitate this and future prioritization exercises. Many other countries (e.g. Germany^28^, USA^29^) currently focus on prioritizing people living in care homes and those with Down Syndrome for vaccination, and should consider broadening this category to include other people with learning disability. Raised risks seen among those under 16 years suggest that vaccination in this age group warrants further consideration. Besides vaccination, efforts should continue to protect people with learning disability from COVID-19 adverse outcomes, including through consideration of non-pharmacological interventions such as shielding, and ensuring adequate support to obtain prompt access to testing for Covid-19 and to appropriate health care.

### Future Research

The ONS data shows that people with disabilities in general are at higher risk of COVID-19 mortality^30^, but this has not yet been explored through the clinical databases, in part because of the complexity of generating codelists for these broad range of conditions. The example of the Learning Disability register data has, however, highlighted the importance of public health surveillance and the need to develop indicators for disability. More research is warranted on the excess COVID-19 risks among people with Down Syndrome. Cerebral palsy includes people with a broad range of conditions and severity, and a deeper exploration or COVID-19 risk for this group is warranted.

### Conclusion

People with learning disabilities have markedly increased risks of hospitalisation and death from COVID-19. Ensuring prompt access to Covid-19 testing and health care and consideration of prioritisation for COVID-19 vaccination and other targeted preventive measures are warranted.

## Data Availability

All data were linked, stored and analysed securely within the OpenSAFELY platform https://opensafely.org/. Data include pseudonymized data such as coded diagnoses, medications and physiological parameters. No free text data are included. All code is shared openly for review and re-use under MIT open license (https://github.com/opensafely/absolute-risks-covid-research). Detailed pseudonymised patient data is potentially re-identifiable and therefore not shared. We rapidly delivered the OpenSAFELY data analysis platform without prior funding to deliver timely analyses on urgent research questions in the context of the global Covid-19 health emergency: now that the platform is established we are developing a formal process for external users to request access in collaboration with NHS England; details of this process will be published shortly on OpenSAFELY.org.

## Administrative

## Acknowledgements

This work uses data provided by patients and collected by the NHS as part of their care and support. We are very grateful for all the support received from the TPP Technical Operations team throughout this work, and for generous assistance from the information governance and database teams at NHS England / NHSX.

## Data sharing

All data were linked, stored and analysed securely within the OpenSAFELY platform https://opensafely.org/. Data include pseudonymized data such as coded diagnoses, medications and physiological parameters. No free text data are included. All code is shared openly for review and re-use under MIT open license (***https://github.com/opensafely/absolute-risks-covid-research***). Detailed pseudonymised patient data is potentially re-identifiable and therefore not shared. We rapidly delivered the OpenSAFELY data analysis platform without prior funding to deliver timely analyses on urgent research questions in the context of the global Covid-19 health emergency: now that the platform is established we are developing a formal process for external users to request access in collaboration with NHS England; details of this process will be published shortly on OpenSAFELY.org.

## Information governance and ethical approval

NHS England is the data controller; TPP is the data processor; and the key researchers on OpenSAFELY are acting on behalf of NHS England. This implementation of OpenSAFELY is hosted within the TPP environment which is accredited to the ISO 27001 information security standard and is NHS IG Toolkit compliant;^31,32^ patient data has been pseudonymised for analysis and linkage using industry standard cryptographic hashing techniques; all pseudonymised datasets transmitted for linkage onto OpenSAFELY are encrypted; access to the platform is via a virtual private network (VPN) connection, restricted to a small group of researchers; the researchers hold contracts with NHS England and only access the platform to initiate database queries and statistical models; all database activity is logged; only aggregate statistical outputs leave the platform environment following best practice for anonymisation of results such as statistical disclosure control for low cell counts.^33^ The OpenSAFELY research platform adheres to the obligations of the UK General Data Protection Regulation (GDPR) and the Data Protection Act 2018. In March 2020, the Secretary of State for Health and Social Care used powers under the UK Health Service (Control of Patient Information) Regulations 2002 (COPI) to require organisations to process confidential patient information for the purposes of protecting public health, providing healthcare services to the public and monitoring and managing the COVID-19 outbreak and incidents of exposure; this sets aside the requirement for patient consent.^34^ Taken together, these provide the legal bases to link patient datasets on the OpenSAFELY platform. GP practices, from which the primary care data are obtained, are required to share relevant health information to support the public health response to the pandemic, and have been informed of the OpenSAFELY analytics platform.

This study was approved by the Health Research Authority (REC reference 20/LO/0651) and by the LSHTM Ethics Board (reference 21863).

## Guarantor

BG is guarantor.

## Contributorship

B.G. conceived the platform and the approach; B.G. and L.S. led the project overall and are guarantors. Contributions are as follows: analysis, E.W., H.I.M., H.K., S.E.; data curation, C.B., J.P., J.C., S.H., S.B., D.E., P.I. and C.E.M.; analysis, E.J.W., K.B., A.J.W. and C.E.M.; funding acquisition, B.G. and L.S.; information governance, A.M., B.G., C.B. and J.P.; methodology, E.W., H.I.M., H.K., S.E, K.B., A.J.W., B.G., L.S., C.B., J.P., J.C., S.H., S.B., D.E., P.I., C.E.M.; disease category conceptualization and codelists, C.E.M., A.J.W., P.I., S.B., D.E., C.B., J.C., J.P., S.H., H.J.C., K.B., S.B., A.M., B.M., L.T., I.J.D., H.I.M., R.M. and H.F.; ethics approval, H.J.C., E.J.W., L.S. and B.G.; project administration, C.E.M., H.J.C., C.B., S.B., A.M., L.S. and B.G.; resources, B.G., L.S. and F.H.; software, S.B., D.E., P.I., A.J.W., C.E.M., S.D., C.B., F.H., J.C. and S.H.; supervision, B.G., L.S. and S.B.; writing (original draft), E.J.W., H.K., H.I.M, S.E. All authors were involved in design and conceptual development and reviewed and approved the final manuscript.

All authors contributed to and approved the final manuscript.

