## Supplementary Appendix for "OpenSAFELY: Risks of COVID-19 hospital admission and death for people with learning disabilities - a cohort study"

The OpenSAFELY Collaborative, Elizabeth J Williamson^1^*, Helen I McDonald^1^*, Krishnan Bhaskaran^1^, Alex J Walker^2^, Sebastian Bacon^2^, Simon Davy^2^, Anna Schultze^1^, Laurie Tomlinson^1^, Chris Bates^3^, Mary Ramsay^4^, Helen J Curtis^2^, Harriet Forbes^1^, Kevin Wing^1^, Caroline Minassian^1^, John Tazare^1^, Caroline E Morton^2^, Emily Nightingale^1^, Amir Mehrkar^2^, Dave Evans^2^, Peter Inglesby^2^, Brian MacKenna^2^, Jonathan Cockburn^3^, Christopher T Rentsch^1^, Rohini Mathur^1^, Angel YS Wong^1^, Rosalind M Eggo^1^, Will Hulme^2^, Richard Croker, John Parry^3^, Frank Hester^3^, Sam Harper^3^, Ian J Douglas^1^, Stephen JW Evans^1^, Liam Smeeth^1^†, Ben Goldacre^2^*†, Hannah Kuper^1^†

† Joint principal investigators; * joint first authors

^1^ London School of Hygiene and Tropical Medicine, Keppel Street, London WC1E 7HT

^2^ The DataLab, Nuffield Department of Primary Care Health Sciences, University of Oxford, OX26GG

^3^ TPP, TPP House, 129 Low Lane, Horsforth, Leeds, LS18 5PX

^4^ National Infection Service, Public Health England, London.

*Corresponding

Table A1a. Estimated hazard ratio for COVID-19 related hospital admissions in wave 1 (1 March 2020 – 31 Aug 2020) in adults 16 and over

|  |  | *Estimated Hazard Ratios (95% confidence intervals)* | | | | |
| --- | --- | --- | --- | --- | --- | --- |
| *Exposure category* | *Events* | *Confounders* | *Confounders*  *+ IMD* | *Confounders +*  *residential care* | *Confounders + comorbidities* | *All* |
| LDR |  |  |  |  |  |  |
| No | 29,635 |  |  |  |  |  |
| Yes | 538 | 5.33 (4.88, 5.83) | 4.84 (4.43, 5.3) | 4.72 (4.29, 5.2) | 4.65 (4.25, 5.09) | 3.93 (3.58, 4.32) |
| by severity: |  |  |  |  |  |  |
| Mild-moderate | 391 | 4.76 (4.32, 5.25) | 4.3 (3.89, 4.74) | 4.37 (3.93, 4.86) | 4.09 (3.7, 4.51) | 3.56 (3.21, 3.95) |
| Severe-profound | 147 | 7.8 (6.47, 9.4) | 7.33 (6.08, 8.82) | 6.45 (5.34, 7.8) | 7.33 (6.07, 8.84) | 5.94 (4.92, 7.16) |
| by residential care status: |  |  |  |  |  |  |
| Not in residential care | 438 | 4.89 (4.46, 5.37) | 4.44 (4.04, 4.87) |  | 4.23 (3.86, 4.65) |  |
| In residential care | 100 | 8.77 (6.8, 11.3) | 8.12 (6.29, 10.48) |  | 8.14 (6.3, 10.51) |  |
| Down's syndrome |  |  |  |  |  |  |
| No | 30,098 |  |  |  |  |  |
| Yes | 75 | 10.56 (8.45, 13.2) | 10.21 (8.17, 12.76) | 6.33 (4.87, 8.23) | 7.21 (5.76, 9.02) | 4.64 (3.58, 6.02) |
| Cerebral Palsy |  |  |  |  |  |  |
| No | 30,076 |  |  |  |  |  |
| Yes | 97 | 4.93 (3.84, 6.34) | 4.64 (3.62, 5.94) | 3.74 (2.91, 4.79) | 4.83 (3.77, 6.2) | 3.78 (2.97, 4.83) |
| Combined grouping |  |  |  |  |  |  |
| None | 29,589 |  |  |  |  |  |
| DS but not LDR | [REDACTED] | 3.92 (1.22, 12.57) | 3.82 (1.19, 12.24) | 3.9 (1.22, 12.47) | 3.56 (1.1, 11.5) | 3.46 (1.07, 11.2) |
| DS and LDR | [REDACTED] | 11.57 (9.2, 14.56) | 11.17 (8.88, 14.07) | 9.96 (7.89, 12.56) | 7.69 (6.1, 9.69) | 6.55 (5.17, 8.29) |
| CP but not LDR | [REDACTED] | 3.39 (2.42, 4.75) | 3.19 (2.28, 4.46) | 3.37 (2.4, 4.72) | 3.34 (2.38, 4.69) | 3.21 (2.29, 4.49) |
| CP and LDR | [REDACTED] | 8.22 (6, 11.27) | 7.72 (5.65, 10.54) | 7.04 (5.17, 9.58) | 7.97 (5.85, 10.85) | 6.62 (4.89, 8.95) |
| LDR with no DS or CP | [REDACTED] | 4.69 (4.26, 5.17) | 4.24 (3.84, 4.67) | 4.23 (3.8, 4.7) | 4.15 (3.77, 4.58) | 3.56 (3.21, 3.95) |

LDR Learning Disability Register; DS Down’s syndrome; CP Cerebral Palsy

Table A1b. Estimated hazard ratio for COVID-19 related death in wave 1 (1 March 2020 – 31 Aug 2020) in adults 16 and over

|  |  | *Estimated Hazard Ratios (95% confidence intervals)* | | | | |
| --- | --- | --- | --- | --- | --- | --- |
| *Exposure category* | *Events* | *Confounders* | *Confounders*  *+ IMD* | *Confounders +*  *residential care* | *Confounders + comorbidities* | *All* |
| LDR |  |  |  |  |  |  |
| No | 13,698 |  |  |  |  |  |
| Yes | 221 | 8.18 (7.12, 9.39) | 7.28 (6.33, 8.38) | 6.96 (5.84, 8.28) | 6.66 (5.78, 7.67) | 5.57 (4.75, 6.52) |
| by severity: |  |  |  |  |  |  |
| Mild-moderate | 158 | 7.1 (6.08, 8.3) | 6.29 (5.37, 7.36) | 6.31 (5.25, 7.58) | 5.72 (4.87, 6.7) | 4.92 (4.14, 5.85) |
| Severe-profound | 63 | 13.21 (9.99, 17.48) | 12.09 (9.14, 15.99) | 10.36 (7.53, 14.24) | 11.35 (8.55, 15.08) | 9.25 (6.85, 12.51) |
| by residential care status: |  |  |  |  |  |  |
| Not in residential care | 182 | 7.82 (6.77, 9.02) | 6.94 (6, 8.02) |  | 6.38 (5.51, 7.39) |  |
| In residential care | 39 | 10.42 (6.88, 15.78) | 9.47 (6.26, 14.35) |  | 8.33 (5.45, 12.73) |  |
| Down's syndrome |  |  |  |  |  |  |
| No | 13,878 |  |  |  |  |  |
| Yes | 41 | 35.96 (26.38, 49.02) | 33.97 (24.91, 46.31) | 18.71 (12.74, 27.47) | 16.18 (11.82, 22.14) | 10.28 (7.15, 14.8) |
| Cerebral Palsy |  |  |  |  |  |  |
| No | 13,889 |  |  |  |  |  |
| Yes | 30 | 5.86 (4.14, 8.3) | 5.46 (3.85, 7.74) | 4.15 (2.82, 6.11) | 5.73 (4.03, 8.15) | 4.6 (3.2, 6.62) |
| Combined grouping |  |  |  |  |  |  |
| None | 13,682 |  |  |  |  |  |
| DS but not LDR | [REDACTED] | 13.45 (3.23, 56.04) | 12.8 (3.06, 53.55) | 13.39 (3.22, 55.59) | 13.08 (3.09, 55.3) | 12.48 (2.94, 52.9) |
| DS and LDR | [REDACTED] | 39.8 (28.87, 54.88) | 37.57 (27.24, 51.81) | 32.38 (22.94, 45.68) | 16.6 (11.95, 23.07) | 14.06 (9.98, 19.81) |
| CP but not LDR | [REDACTED] | 3.82 (2.3, 6.36) | 3.58 (2.15, 5.95) | 3.8 (2.28, 6.31) | 3.73 (2.23, 6.23) | 3.55 (2.12, 5.92) |
| CP and LDR | [REDACTED] | 12.02 (7.47, 19.33) | 11.1 (6.9, 17.86) | 9.86 (5.95, 16.33) | 11.89 (7.42, 19.04) | 9.97 (6.12, 16.26) |
| LDR with no DS or CP | [REDACTED] | 6.78 (5.82, 7.89) | 6.01 (5.16, 7) | 5.92 (4.93, 7.1) | 5.67 (4.86, 6.62) | 4.83 (4.08, 5.72) |

LDR Learning Disability Register; DS Down’s syndrome; CP Cerebral Palsy

Table A1c. Estimated hazard ratio for COVID-19 related hospital admissions in wave 2 (1 September 2020 – 31 November 2020) in adults 16 and over

|  |  | *Estimated Hazard Ratios (95% confidence intervals)* | | | | |
| --- | --- | --- | --- | --- | --- | --- |
| *Exposure category* | *Events* | *Confounders* | *Confounders*  *+ IMD* | *Confounders +*  *residential care* | *Confounders + comorbidities* | *All* |
| LDR |  |  |  |  |  |  |
| No | 27,228 |  |  |  |  |  |
| Yes | 383 | 3.64 (3.28, 4.03) | 3.29 (2.96, 3.65) | 3.35 (2.99, 3.75) | 3.23 (2.92, 3.59) | 2.81 (2.51, 3.14) |
| by severity: |  |  |  |  |  |  |
| Mild-moderate | 281 | 3.27 (2.91, 3.67) | 2.93 (2.6, 3.3) | 3.09 (2.73, 3.5) | 2.86 (2.54, 3.21) | 2.53 (2.24, 2.87) |
| Severe-profound | 102 | 5.27 (4.31, 6.44) | 4.92 (4.03, 6.02) | 4.63 (3.76, 5.69) | 5.1 (4.18, 6.21) | 4.32 (3.53, 5.29) |
| by residential care status: |  |  |  |  |  |  |
| Not in residential care | 318 | 3.36 (2.99, 3.76) | 3.03 (2.69, 3.41) |  | 2.96 (2.64, 3.31) |  |
| In residential care | 65 | 6.14 (4.58, 8.22) | 5.66 (4.23, 7.57) |  | 5.95 (4.46, 7.96) |  |
| Down's syndrome |  |  |  |  |  |  |
| No | 27,558 |  |  |  |  |  |
| Yes | 53 | 7.22 (5.52, 9.46) | 7.01 (5.36, 9.17) | 5.35 (4.05, 7.09) | 5.4 (4.12, 7.06) | 4.17 (3.16, 5.5) |
| Cerebral Palsy |  |  |  |  |  |  |
| No | 27,536 |  |  |  |  |  |
| Yes | 75 | 3.7 (2.93, 4.69) | 3.46 (2.73, 4.4) | 3.13 (2.42, 4.04) | 3.63 (2.85, 4.62) | 3.05 (2.36, 3.95) |
| Combined grouping |  |  |  |  |  |  |
| None | 27,193 |  |  |  |  |  |
| DS but not LDR | [REDACTED] | 2.43 (.58, 10.13) | 2.38 (.57, 9.9) | 2.43 (.58, 10.1) | 2.18 (.52, 9.08) | 2.14 (.51, 8.91) |
| DS and LDR | [REDACTED] | 7.85 (5.98, 10.31) | 7.61 (5.79, 9.98) | 7.14 (5.45, 9.34) | 5.75 (4.38, 7.55) | 5.19 (3.96, 6.79) |
| CP but not LDR | [REDACTED] | 2.56 (1.83, 3.58) | 2.4 (1.71, 3.36) | 2.55 (1.82, 3.56) | 2.52 (1.8, 3.54) | 2.4 (1.7, 3.37) |
| CP and LDR | [REDACTED] | 5.94 (4.4, 8.03) | 5.55 (4.1, 7.52) | 5.34 (3.92, 7.28) | 5.79 (4.27, 7.84) | 5 (3.67, 6.83) |
| LDR with no DS or CP | [REDACTED] | 3.17 (2.83, 3.56) | 2.85 (2.53, 3.2) | 2.96 (2.61, 3.35) | 2.85 (2.54, 3.19) | 2.49 (2.2, 2.81) |

LDR Learning Disability Register; DS Down’s syndrome; CP Cerebral Palsy

Table A1d. Estimated hazard ratio for COVID-19 related death in wave 2 (1 September 2020 – January 2021) in adults 16 and over

|  |  | *Estimated Hazard Ratios (95% confidence intervals)* | | | | |
| --- | --- | --- | --- | --- | --- | --- |
| *Exposure category* | *Events* | *Confounders* | *Confounders*  *+ IMD* | *Confounders +*  *residential care* | *Confounders + comorbidities* | *All* |
| LDR |  |  |  |  |  |  |
| No | 17,673 |  |  |  |  |  |
| Yes | 260 | 7.39 (6.52, 8.38) | 6.57 (5.77, 7.47) | 6.88 (5.99, 7.89) | 6.47 (5.7, 7.34) | 5.7 (4.98, 6.53) |
| by severity: |  |  |  |  |  |  |
| Mild-moderate | 179 | 6.2 (5.37, 7.17) | 5.48 (4.73, 6.34) | 5.94 (5.12, 6.9) | 5.32 (4.6, 6.16) | 4.79 (4.13, 5.57) |
| Severe-profound | 81 | 12.81 (10.25, 16) | 11.71 (9.35, 14.67) | 11.72 (9.15, 15.01) | 12.37 (9.93, 15.41) | 11.14 (8.76, 14.16) |
| by residential care status: |  |  |  |  |  |  |
| Not in residential care | 211 | 6.92 (6.04, 7.94) | 6.13 (5.32, 7.06) | 6.93 (6.04, 7.94) | 6.04 (5.26, 6.94) |  |
| In residential care | 49 | 10.41 (7.51, 14.43) | 9.44 (6.79, 13.11) | 5.01 (1.97, 12.77) | 9.3 (6.76, 12.8) |  |
| Down's syndrome |  |  |  |  |  |  |
| No | 17,868 |  |  |  |  |  |
| Yes | 65 | 42.91 (33.54, 54.9) | 40.6 (31.72, 51.96) | 29.77 (22, 40.28) | 24.6 (19.27, 31.41) | 18.52 (13.8, 24.87) |
| Cerebral Palsy |  |  |  |  |  |  |
| No | 17,902 |  |  |  |  |  |
| Yes | 31 | 4.54 (3.19, 6.47) | 4.23 (2.97, 6.02) | 3.46 (2.37, 5.06) | 4.63 (3.25, 6.6) | 3.74 (2.59, 5.41) |
| Combined grouping |  |  |  |  |  |  |
| None | 17,660 |  |  |  |  |  |
| DS but not LDR | [REDACTED] |  |  |  |  |  |
| DS and LDR | [REDACTED] | 50.43 (39.47, 64.43) | 47.73 (37.37, 60.94) | 46.85 (36.35, 60.38) | 27.06 (21.13, 34.64) | 25.36 (19.6, 32.81) |
| CP but not LDR | [REDACTED] | 2.67 (1.58, 4.5) | 2.5 (1.48, 4.22) | 2.67 (1.58, 4.49) | 2.69 (1.6, 4.54) | 2.56 (1.52, 4.33) |
| CP and LDR | [REDACTED] | 10.01 (6.18, 16.21) | 9.23 (5.71, 14.93) | 9.25 (5.65, 15.15) | 10.6 (6.56, 17.14) | 9.56 (5.86, 15.58) |
| LDR with no DS or CP | [REDACTED] | 5.56 (4.76, 6.5) | 4.92 (4.19, 5.77) | 5.29 (4.5, 6.22) | 4.95 (4.24, 5.78) | 4.45 (3.79, 5.23) |

LDR Learning Disability Register; DS Down’s syndrome; CP Cerebral Palsy

Table A2a. Estimated hazard ratio for COVID-19 related hospital admissions in wave 1 (1 March – 31^st^ August 2020) in children under 16

|  | *Estimated Hazard Ratios (95% confidence intervals)* | | | | |
| --- | --- | --- | --- | --- | --- |
| *Exposure category* | *Confounders* | *Confounders*  *+ IMD* | *Confounders +*  *residential care* | *Confounders + comorbidities* | *All* |
| LDR |  |  |  |  |  |
| No |  |  |  |  |  |
| Yes | 5.13 (1.88, 14) | 5.06 (1.85, 13.81) | 5.15 (1.89, 14.06) | 5.16 (1.89, 14.09) | 5.11 (1.87, 13.95) |
| by severity: |  |  |  |  |  |
| Mild-moderate | 3.1 (0.76, 12.59) | 3.06 (.75, 12.4) | 3.11 (.77, 12.62) | 3.12 (.77, 12.66) | 3.08 (.76, 12.51) |
| Severe-profound | 14.92 (3.63, 61.3) | 14.76 (3.59, 60.66) | 15.11 (3.68, 62.11) | 15.04 (3.66, 61.85) | 15.07 (3.67, 62) |
| by residential care status: |  |  |  |  |  |
| Not in residential care | 5.15 (1.89, 14.06) | 5.08 (1.86, 13.87) | 5.18 (1.9, 14.15) | 5.11 (1.87, 13.95) | 5.15 (1.89, 14.06) |
| In residential care | * |  |  |  |  |
| Down's syndrome |  |  |  |  |  |
| No |  |  |  |  |  |
| Yes | 6.85 (1.69, 27.71) | 6.83 (1.69, 27.61) | 6.85 (1.69, 27.73) | 6.88 (1.7, 27.81) | 6.86 (1.7, 27.73) |
| Cerebral Palsy |  |  |  |  |  |
| No |  |  |  |  |  |
| Yes | 10.75 (4.25, 27.22) | 10.71 (4.23, 27.13) | 10.77 (4.25, 27.26) | 10.77 (4.25, 27.26) | 10.74 (4.24, 27.2) |
| Combined grouping |  |  |  |  |  |
| None |  |  |  |  |  |
| DS but not LDR | 4.82 (.67, 34.46) | 4.81 (.67, 34.37) | 4.82 (.67, 34.47) | 4.82 (.67, 34.48) | 4.81 (.67, 34.4) |
| DS and LDR | 12.66 (1.78, 89.92) | 12.6 (1.78, 89.43) | 12.68 (1.79, 90.03) | 12.8 (1.8, 90.98) | 12.76 (1.8, 90.62) |
| CP but not LDR | 9.2 (3.18, 26.65) | 9.17 (3.16, 26.57) | 9.2 (3.18, 26.65) | 9.21 (3.18, 26.68) | 9.18 (3.17, 26.61) |
| CP and LDR | 24.79 (3.49, 176.18) | 24.61 (3.46, 174.79) | 25.12 (3.53, 178.59) | 24.87 (3.5, 176.71) | 24.99 (3.52, 177.54) |
| LDR with no DS or CP | 3.09 (.76, 12.59) | 3.04 (.74, 12.41) | 3.1 (.76, 12.64) | 3.1 (.76, 12.66) | 3.07 (.75, 12.54) |

LDR Learning Disability Register; DS Down’s syndrome; CP Cerebral Palsy

Table A2b. Estimated hazard ratio for COVID-19 related hospital admissions in wave 2 (1 September– December 2020) in children under 16

|  | *Estimated Hazard Ratios (95% confidence intervals)* | | | | |
| --- | --- | --- | --- | --- | --- |
| *Exposure category* | *Confounders* | *Confounders*  *+ IMD* | *Confounders +*  *residential care* | *Confounders + comorbidities* | *All* |
| LDR |  |  |  |  |  |
| No |  |  |  |  |  |
| Yes | 5.13 (1.88, 14) | 5.06 (1.85, 13.81) | 5.15 (1.89, 14.06) | 5.16 (1.89, 14.09) | 5.11 (1.87, 13.95) |
| by severity: |  |  |  |  |  |
| Mild-moderate | 3.1 (0.76, 12.59) | 3.06 (.75, 12.4) | 3.11 (.77, 12.62) | 3.12 (.77, 12.66) | 3.08 (.76, 12.51) |
| Severe-profound | 14.92 (3.63, 61.3) | 14.76 (3.59, 60.66) | 15.11 (3.68, 62.11) | 15.04 (3.66, 61.85) | 15.07 (3.67, 62) |
| by residential care status: |  |  |  |  |  |
| Not in residential care | 5.15 (1.89, 14.06) | 5.08 (1.86, 13.87) | 5.18 (1.9, 14.15) | 5.11 (1.87, 13.95) | 5.15 (1.89, 14.06) |
| In residential care | * |  |  |  |  |
| Down's syndrome |  |  |  |  |  |
| No |  |  |  |  |  |
| Yes | 6.85 (1.69, 27.71) | 6.83 (1.69, 27.61) | 6.85 (1.69, 27.73) | 6.88 (1.7, 27.81) | 6.86 (1.7, 27.73) |
| Cerebral Palsy |  |  |  |  |  |
| No |  |  |  |  |  |
| Yes | 10.75 (4.25, 27.22) | 10.71 (4.23, 27.13) | 10.77 (4.25, 27.26) | 10.77 (4.25, 27.26) | 10.74 (4.24, 27.2) |
| Combined grouping |  |  |  |  |  |
| None |  |  |  |  |  |
| DS but not LDR | 4.82 (.67, 34.46) | 4.81 (.67, 34.37) | 4.82 (.67, 34.47) | 4.82 (.67, 34.48) | 4.81 (.67, 34.4) |
| DS and LDR | 12.66 (1.78, 89.92) | 12.6 (1.78, 89.43) | 12.68 (1.79, 90.03) | 12.8 (1.8, 90.98) | 12.76 (1.8, 90.62) |
| CP but not LDR | 9.2 (3.18, 26.65) | 9.17 (3.16, 26.57) | 9.2 (3.18, 26.65) | 9.21 (3.18, 26.68) | 9.18 (3.17, 26.61) |
| CP and LDR | 24.79 (3.49, 176.18) | 24.61 (3.46, 174.79) | 25.12 (3.53, 178.59) | 24.87 (3.5, 176.71) | 24.99 (3.52, 177.54) |
| LDR with no DS or CP | 3.09 (.76, 12.59) | 3.04 (.74, 12.41) | 3.1 (.76, 12.64) | 3.1 (.76, 12.66) | 3.07 (.75, 12.54) |

LDR Learning Disability Register; DS Down’s syndrome; CP Cerebral Palsy; * insufficient numbers to estimate.

Table A3. Estimated hazard ratio for being on the learning disability register for COVID-19 related outcomes – interactions with age

|  |  | *Estimated Hazard Ratios (95% confidence intervals)* | | | | |
| --- | --- | --- | --- | --- | --- | --- |
| *Outcome* | *Age group* | *Confounders* | *With IMD* | *With Residential care* | *Comorbidities* | *All* |
|  | **Wave 1: 1 March– 31 August 2020** | | | | | |
| COVID-19 related hospital admission | 16-<65 | 5.42 (4.87, 6.03) | 5 (4.49, 5.55) | 4.88 (4.37, 5.44) | 4.54 (4.08, 5.06) | 3.93 (3.53, 4.38) |
|  | 65-<75 | 6.52 (5.43, 7.83) | 5.82 (4.83, 7) | 5.59 (4.63, 6.76) | 5.74 (4.77, 6.9) | 4.66 (3.86, 5.63) |
|  | 75+ | 3.57 (2.81, 4.54) | 3.17 (2.49, 4.03) | 3.04 (2.36, 3.93) | 3.62 (2.84, 4.62) | 2.90 (2.25, 3.75) |
| COVID-19 death | 16-<65 | 12.25 (9.96, 15.07) | 11.11 (9.03, 13.67) | 10.57 (8.46, 13.22) | 9.47 (7.7, 11.65) | 8.13 (6.57, 10.06) |
|  | 65-<75 | 10.5 (8.32, 13.25) | 9.33 (7.39, 11.78) | 8.78 (6.81, 11.32) | 8.03 (6.36, 10.15) | 6.65 (5.21, 8.48) |
|  | 75+ | 4.16 (3.17, 5.45) | 3.67 (2.79, 4.82) | 3.44 (2.52, 4.7) | 3.66 (2.77, 4.83) | 2.99 (2.22, 4.04) |
|  | **Wave 2: 1 September 2020 – 31 December 2020 (admissions) or 8 February 2021 (death)** | | | | | |
| COVID-19 related hospital admission | 16-<65 | 3.38 (2.98, 3.83) | 3.1 (2.73, 3.51) | 3.15 (2.77, 3.59) | 2.88 (2.54, 3.26) | 2.56 (2.26, 2.91) |
|  | 65-<75 | 4.85 (3.95, 5.96) | 4.27 (3.47, 5.25) | 4.38 (3.53, 5.45) | 4.4 (3.57, 5.41) | 3.70 (2.98, 4.59) |
|  | 75+ | 3.36 (2.57, 4.38) | 2.95 (2.25, 3.87) | 3.02 (2.27, 4.01) | 3.57 (2.74, 4.64) | 2.93 (2.21, 3.89) |
| COVID-19 death | 16-<65 | 11.45 (9.48, 13.82) | 10.35 (8.56, 12.53) | 10.68 (8.77, 13.01) | 9.24 (7.66, 11.15) | 8.29 (6.83, 10.06) |
|  | 65-<75 | 8.01 (6.4, 10.02) | 7.1 (5.69, 8.86) | 7.37 (5.87, 9.26) | 6.67 (5.34, 8.33) | 5.86 (4.68, 7.33) |
|  | 75+ | 3.99 (3.05, 5.21) | 3.5 (2.66, 4.61) | 3.65 (2.76, 4.81) | 3.84 (2.94, 5.02) | 3.29 (2.49, 4.35) |

Table A4. Estimated hazard ratio for being on the learning disability register for COVID-19 related hospital admission, after excluding those prioritised for vaccination through age or comorbidity in groups 1-6 of Phase I vaccination

|  |  | *Estimated Hazard Ratios (95% confidence intervals)* | | |
| --- | --- | --- | --- | --- |
| *Exposure* | *Exposure category* | *Confounders* | *With IMD* | *With Residential care* |
| Learning disability register | No |  |  |  |
|  | Yes | 6.66 (1.57, 28.22) | 6.61 (1.54, 28.27) | 6.98 (1.65, 29.51) |

Table A5a. Estimated hazard ratio for non-COVID-19 related death in wave 1 (1 March – 31 August 2020) in adults 16 and over

|  |  | *Estimated Hazard Ratios (95% confidence intervals)* | | | | |
| --- | --- | --- | --- | --- | --- | --- |
| *Exposure category* | *Events* | *Confounders* | *Confounders*  *+ IMD* | *Confounders +*  *residential care* | *Confounders + comorbidities* | *All* |
| LDR |  |  |  |  |  |  |
| No | 69207 |  |  |  |  |  |
| Yes | 596 | 3.71 (3.42, 4.03) | 3.38 (3.1, 3.68) | 3.32 (3, 3.69) | 3.44 (3.17, 3.73) | 2.99 (2.72, 3.29) |
| by severity: |  |  |  |  |  |  |
| Mild-moderate | 424 | 3.2 (2.91, 3.52) | 2.9 (2.63, 3.2) | 2.97 (2.66, 3.31) | 2.94 (2.67, 3.23) | 2.61 (2.36, 2.9) |
| Severe-profound | 172 | 6.1 (5.24, 7.1) | 5.7 (4.89, 6.64) | 5.19 (4.32, 6.24) | 5.96 (5.11, 6.95) | 5.14 (4.33, 6.11) |
| by residential care status: |  |  |  |  |  |  |
| Not in residential care | 498 | 3.61 (3.3, 3.94) | 3.28 (2.99, 3.59) |  | 6.04 (5.26, 6.94) |  |
| In residential care | 98 | 4.35 (3.54, 5.35) | 4.04 (3.27, 4.98) |  | 9.3 (6.76, 12.8) |  |
| Down's syndrome |  |  |  |  |  |  |
| No | 69703 |  |  |  |  |  |
| Yes | 100 | 12.31 (9.97, 15.2) | 11.83 (9.56, 14.65) | 8.7 (6.71, 11.29) | 8.03 (6.45, 10.01) | 6.19 (4.79, 8.01) |
| Cerebral Palsy |  |  |  |  |  |  |
| No | 69705 |  |  |  |  |  |
| Yes | 98 | 3.21 (2.6, 3.95) | 3.03 (2.45, 3.75) | 2.73 (2.18, 3.41) | 3.2 (2.6, 3.94) | 2.79 (2.25, 3.46) |
| Combined grouping |  |  |  |  |  |  |
| None | 69157 |  |  |  |  |  |
| DS but not LDR | [REDACTED] | 2.06 (.5, 8.5) | 1.98 (.48, 8.19) | 2.05 (.5, 8.47) | 2.07 (.5, 8.59) | 1.99 (.48, 8.27) |
| DS and LDR | [REDACTED] | 13.82 (11.06, 17.28) | 13.28 (10.6, 16.63) | 12.05 (9.42, 15.42) | 8.61 (6.8, 10.9) | 7.72 (6, 9.93) |
| CP but not LDR | [REDACTED] | 2.22 (1.64, 3.02) | 2.1 (1.54, 2.87) | 2.21 (1.63, 3) | 2.19 (1.63, 2.95) | 2.1 (1.56, 2.83) |
| CP and LDR | [REDACTED] | 5.8 (4.42, 7.63) | 5.45 (4.15, 7.17) | 5.08 (3.83, 6.75) | 5.96 (4.54, 7.84) | 5.23 (3.95, 6.92) |
| LDR with no DS or CP | [REDACTED] | 3.1 (2.83, 3.4) | 2.81 (2.57, 3.08) | 2.83 (2.55, 3.15) | 2.93 (2.67, 3.21) | 2.58 (2.34, 2.85) |

LDR Learning Disability Register; DS Down’s syndrome; CP Cerebral Palsy

Table A5b. Estimated hazard ratio for non-COVID-19 related death in wave 2 (1 September 2020 – January 2021) in adults 16 and over

|  |  | *Estimated Hazard Ratios (95% confidence intervals)* | | | | |
| --- | --- | --- | --- | --- | --- | --- |
| *Exposure category* | *Events* | *Confounders* | *Confounders*  *+ IMD* | *Confounders +*  *residential care* | *Confounders + comorbidities* | *All* |
| LDR |  |  |  |  |  |  |
| No | 53701 |  |  |  |  |  |
| Yes | 470 | 3.91 (3.58, 4.27) | 3.59 (3.29, 3.93) | 3.6 (3.24, 4) | 3.67 (3.35, 4.01) | 3.25 (2.95, 3.59) |
| by severity: |  |  |  |  |  |  |
| Mild-moderate | 353 | 3.56 (3.22, 3.94) | 3.25 (2.94, 3.6) | 3.36 (3, 3.76) | 3.3 (2.98, 3.66) | 2.97 (2.67, 3.32) |
| Severe-profound | 117 | 5.57 (4.65, 6.66) | 5.23 (4.37, 6.26) | 4.91 (4.01, 6.01) | 5.51 (4.61, 6.59) | 4.84 (3.99, 5.88) |
| by residential care status: |  |  |  |  |  |  |
| Not in residential care | 397 | 3.83 (3.48, 4.21) | 3.5 (3.19, 3.86) |  | 3.58 (3.25, 3.94) |  |
| In residential care | 73 | 4.45 (3.51, 5.65) | 4.16 (3.27, 5.29) | 4.22 (3.32, 5.36) | 9.3 (6.76, 12.8) |  |
| Down's syndrome |  |  |  |  |  |  |
| No | 54116 |  |  |  |  |  |
| Yes | 55 | 9.52 (7.19, 12.6) | 9.17 (6.92, 12.13) | 6.6 (4.97, 8.77) | 6.38 (4.84, 8.41) | 4.66 (3.49, 6.22) |
| Cerebral Palsy |  |  |  |  |  |  |
| No | 54111 |  |  |  |  |  |
| Yes | 60 | 2.61 (2.04, 3.34) | 2.48 (1.94, 3.17) | 2.21 (1.7, 2.86) | 2.68 (2.08, 3.44) | 2.3 (1.77, 2.98) |
| Combined grouping |  |  |  |  |  |  |
| None | 53672 |  |  |  |  |  |
| DS but not LDR | [REDACTED] | * |  |  |  |  |
| DS and LDR | [REDACTED] | 11.07 (8.41, 14.56) | 10.66 (8.1, 14.02) | 9.95 (7.56, 13.11) | 7.06 (5.37, 9.28) | 6.39 (4.85, 8.43) |
| CP but not LDR | [REDACTED] | 1.78 (1.25, 2.53) | 1.69 (1.19, 2.4) | 1.77 (1.24, 2.52) | 1.8 (1.27, 2.56) | 1.73 (1.22, 2.46) |
| CP and LDR | [REDACTED] | 4.87 (3.43, 6.91) | 4.58 (3.22, 6.52) | 4.37 (3.02, 6.31) | 5.11 (3.58, 7.3) | 4.53 (3.13, 6.55) |
| LDR with no DS or CP | [REDACTED] | 3.53 (3.21, 3.89) | 3.23 (2.93, 3.56) | 3.29 (2.94, 3.68) | 3.36 (3.05, 3.71) | 3 (2.7, 3.34) |

LDR Learning Disability Register; DS Down’s syndrome; CP Cerebral Palsy; * insufficient numbers to estimate.

Table A1. Codelists used to define variables used in the analysis

| Variable | Notes | Codelist |
| --- | --- | --- |
| On the learning disability register | Plus any individuals identified as severe and profound learning disability (all of whom should be on the learning disability register) | <https://codelists.opensafely.org/codelist/opensafely/learning-disabilities/2020-07-06/> |
| Severe and profound learning disability |  | <https://codelists.opensafely.org/codelist/opensafely/severe-and-profound-learning-disability-flags/44ef542a/#full-list> |
| Down’s syndrome |  | <https://codelists.opensafely.org/codelist/opensafely/down-syndrome/15832db6/#full-list> |
| Cerebral Palsy |  | <https://codelists.opensafely.org/codelist/opensafely/cerebral-palsy/1835edac/#full-list> |
| Ethnicity | 5 categories, obtained from 16 (White, South Asian, Black, Mixed, Other) | [https://codelists.opensafely.org/codelist/opensafely/ethnicity](https://codelists.opensafely.org/codelist/opensafely/ethnicity/) |
| C[hronic cardiac disease](https://codelists.opensafely.org/codelist/opensafely/chronic-cardiac-disease/) |  | <https://codelists.opensafely.org/codelist/opensafely/chronic-cardiac-disease/> |
| Atrial Fibrillation | This codelist includes both atrial fibrillation and atrial flutter | <https://codelists.opensafely.org/codelist/opensafely/atrial-fibrillation-or-flutter/2020-07-30/> |
| Prior deep vein thrombosis / pulmonary embolism |  | <https://codelists.opensafely.org/codelist/opensafely/venous-thromboembolic-disease/2020-09-14/> |
| D[iabetes](https://codelists.opensafely.org/codelist/opensafely/diabetes/) | Combined with Hba1c measure within 18 months to determine level of control | <https://codelists.opensafely.org/codelist/opensafely/diabetes/> |
| S[troke](https://codelists.opensafely.org/codelist/opensafely/stroke-updated/) and transient ischaemic attack |  | <https://codelists.opensafely.org/codelist/opensafely/stroke-updated/2020-06-02/>  <https://codelists.opensafely.org/codelist/opensafely/transient-ischaemic-attack/3526e2ac/> |
| Dementia |  | <https://codelists.opensafely.org/codelist/opensafely/dementia-complete/48c76cf8> |
| O[ther neurological conditions](https://codelists.opensafely.org/codelist/opensafely/other-neurological-conditions/) | Will be used only for exclusion of individuals eligible for vaccination in final analysis: will not be adjusted for in models. | <https://codelists.opensafely.org/codelist/opensafely/other-neurological-conditions/> |
| Asthma | Combined with OCS prescriptions in past year to determine severity | <https://codelists.opensafely.org/codelist/opensafely/asthma-diagnosis/> |
| Cystic Fibrosis and associated conditions |  | <https://codelists.opensafely.org/codelist/opensafely/cystic-fibrosis/2020-07-20/> |
| R[espiratory disease other than asthma](https://codelists.opensafely.org/codelist/opensafely/chronic-respiratory-disease/) or cystic fibrosis |  | <https://codelists.opensafely.org/codelist/opensafely/other-chronic-respiratory-disease/2020-07-20/> |
| Non-haematological cancer | Incident diagnosis within the previous year | <https://codelists.opensafely.org/codelist/opensafely/cancer-excluding-lung-and-haematological/> |
| Haematological cancer | Grouped by time since diagnosis (<1 year, 2-<5 years, 5+years) | <https://codelists.opensafely.org/codelist/opensafely/haematological-cancer/> |
| Lung cancer | Incident diagnosis within the previous year, combined with other non-haematological cancer | <https://codelists.opensafely.org/codelist/opensafely/lung-cancer/> |
| L[iver disease](https://codelists.opensafely.org/codelist/opensafely/chronic-liver-disease/) |  | <https://codelists.opensafely.org/codelist/opensafely/chronic-liver-disease/> |
| Kidney dialysis | Used if no kidney transplant since most recent dialysis | <https://codelists.opensafely.org/codelist/opensafely/dialysis/2020-07-16/> |
| Kidney transplant | Combined with non-kidney transplant for transplant indicator. Also used to determine which of dialysis/transplant is most recent. | <https://codelists.opensafely.org/codelist/opensafely/kidney-transplant/2020-07-15/> |
| Organ transplant (other than kidney) | Combined with kidney transplant for transplant indicator. | <https://codelists.opensafely.org/codelist/opensafely/other-organ-transplant/2020-07-15/> |
| A[splenia](https://codelists.opensafely.org/codelist/opensafely/asplenia/) |  | <https://codelists.opensafely.org/codelist/opensafely/asplenia/> <https://codelists.opensafely.org/codelist/opensafely/sickle-cell-disease/> |
| R[heumatoid arthritis, lupus, psoriasis](https://codelists.opensafely.org/codelist/opensafely/ra-sle-psoriasis/) |  | <https://codelists.opensafely.org/codelist/opensafely/ra-sle-psoriasis/> |
| Other immunosuppressive condition | Temporary and aplastic anaemia within last year; HIV and other permanent immunosuppression ever. | <https://codelists.opensafely.org/codelist/opensafely/hiv/2020-07-13/>  <https://codelists.opensafely.org/codelist/opensafely/permanent-immunosuppresion/>  <https://codelists.opensafely.org/codelist/opensafely/aplastic-anaemia/>  <https://codelists.opensafely.org/codelist/opensafely/temporary-immunosuppresion/> |
| Inflammatory bowel disease |  | <https://codelists.opensafely.org/codelist/opensafely/inflammatory-bowel-disease/2020-04-07/> |
| Serious mental illness | Psychosis, schizophrenia and bipolar affective disease | <https://codelists.opensafely.org/codelist/opensafely/psychosis-schizophrenia-bipolar-affective-disease/2020-07-09/> |
